# Genome-wide transcriptome analysis reveals sex-specific biological differences in the early phase of an acute myocardial infarction

**DOI:** 10.1101/2025.02.19.25322579

**Authors:** Aaron Shulkin, Perman Pandal, Eliseo Vazquez, Elizabeth Cortez-Toledo, Kwame Atsina, Tesfaye B. Mersha, Javier E. López

**Affiliations:** Carle Illinois College of Medicine, University of Illinois Urbana-Champaign, Urbana, IL, USA; Cardiovascular Division, Department of Internal Medicine, University of California Davis, Sacramento, CA, USA; Department of Pediatrics, Cincinnati Children’s Hospital Medical Center, University of Cincinnati, Cincinnati, OH, USA; Cardiovascular Research Institute, University of California Davis, Davis, CA, USA

## Abstract

**Background:** Clinical outcomes of acute myocardial infarction (AMI) are known to vary between females and males; however, the nature of this sex dimorphism remains controversial. Most AMI transcriptomic studies have not considered differences between females and males, and combined sexes in their analysis to increase sample size and gain power (canonical approach). Our objective was to (1) use a sex-specific differentially expressed gene meta-analysis (ss-DEGma) in blood and (2) identify sex-specific pathways related to the early phase of AMI.

**Methods:** Gene expression data (7 sets) for sex-combined (canonical) and sex-specific analysis (ss-DEGma) were obtained from the publicly-available GEO database. Datasets from whole blood and peripheral blood cells sampled within 3 days post-AMI were analyzed using GEO2R. The massiR tool identified sex in 72% of samples. The top-ranking DEGs were used to identify significant sex-specific biological pathways in the KEGG database (FDR <0.05).

**Results:** We performed this meta-analysis in 291 women and 452 men and > 20,000 genes (see Table for identified DEGs). Sex-combined DEGs yielded 100 significant KEGG pathways. Sex-specific DEGs yielded 8/61 (13%) additional new pathways not identified by the sex-combined analysis. Sex-combined pathways were predominantly immunological (35%), while male- and female-specific pathways were 43% and 18% immunological, respectively. Proliferative and metabolic pathways were the next most represented pathways in females, which were not present in males at all.

**Conclusion:** We present 8 new sex-specific AMI-related transcriptional pathways not identified in the canonical sex-combined analysis. Furthermore, we find that 53% of pathways identified in the canonical sex-combined analysis are not shared between sexes. This data underscores an urgent need for prospective sex-specific transcriptomic analysis to define the sex-specific biological difference post-AMI.

|  | *Combined*<br>*Sexes 483 AMI*<br>*and*<br>*260 CNTL* | *Women only\*\**<br>*164 AMI and*<br>*127 CNTL* | *Men only\*\**<br>*319 AMI and*<br>*133 CNTL* |
| --- | --- | --- | --- |
| *Number of*<br>*Genes with raw*<br>*p < 0.05* | **4188** | **1074** | **1488** |
| *Number of*<br>*Pathways with*<br>*FDR < 0.05\** | **100** | **14\*\*** | **53\*\*** |

\*Pathways were derived from top 1000 gene for each group i.e. Combined, Men or Women using NetworkAnalyst. \*\* 56 out these 64 pathways were shared with the 100 canonical sex-combined pathways, whereas none were shared between females and males. CNTL=control

## INTRODUCTION

Differences in disease severity or outcomes on the basis of sex is a well-documented phenomenon and seen across several pathologies including cardiovascular disease, asthma, and autoimmunity [1 2]. While cardiovascular deaths are the leading cause of mortality in both men and women, with 931,578 reported in the last year, there are several differences in the prevention, presentation, and management of cardiovascular disease between sexes [3–5]. These can include delays in seeking medical advice, underuse of medical imaging, misinterpretation of signs and symptoms, and many others. For example, the American Heart Association estimates the average age of a male’s first acute myocardial infarction (AMI) is at 65.6 years old versus 72.0 years old for females. According to Mehta et al, more women than men will die (26% of women and 19% men) within a year of a first AMI; more women than men will die (47% of women and 36% of men) within 5 years of first AMI, regardless of age [6]. These sex-specific differences with respect to mortality among AMI patients have been investigated in several studies; however, the causes remain controversial.

It has been theorized that there is a genetic and transcriptomic arm to the manifestation of these sexual dimorphisms seen on the clinical level. Previous studies have demonstrated this theory’s credibility through genome wide transcriptomic and association studies in asthma but very few have been explored for AMIs [2 7]. Gautam et al pioneered sex-specific transcriptomic studies in asthma and developed a pipeline which identified 439 male- and 297 female-specific genes involved in asthma etiology [2]. Their study showcased how sex-specific analyses yield new information, not previously seen in more canonical, sex-combined studies and demonstrate a possible avenue to be informative in the cardiovascular sphere. Dungan et al identified 8 SNPs specific to men and 15 SNPs specific to women implicated in all-cause mortality following acute MI [7]. While this ischemic heart disease genome wide association study (GWAS) failed to meet high significance thresholds, it underpins a crucial idea that cardiac sex dimorphisms can persist beyond modifiable risk factors, and that transcriptomic differences do play a role in clinical manifestations. Together, these studies demonstrate the need for further genomic and transcriptomic characterization of acute MI mortality and the sexual dimorphism that plays out in the hospital.

Understanding how men and women differ post-AMI is key to properly treating patients and improving outcomes. Better characterization of these differences, as well as other relevant clinical treatments, aim to improve the disparities female patients face when treated for myocardial infarction. Previous analyses have yielded important insights to the outcome sexual dimorphism and warrant further investigation into transcriptomic differences between sexes.

Here, we perform a sex-specific analysis to study the differences in gene expression between men and women following AMI. Previously, Zhou et al. published the first ever article studying gene expression differences between male and female patients with AMI [8]. However, this study was limited due to a small sample size and exclusion of some datasets that did not contain gender information, both of which we aimed to overcome during our analysis. The purpose of this study is to identify sex-specific differentially expressed genes (DEGs), and sex-specific pathways previously not characterized in canonical AMI studies.

## METHODS

### Publicly-available transcriptomic data source

We collected microarray expression datasets publicly available from the Gene Expression Omnibus (GEO) database [9] as previously done [2]. Our data pipeline for analysis is depicted in Figure 1. Briefly, we searched the GEO database from May 1, 2011 to December 31, 2018 for data that met the following criteria: (a) dataset must compare humans older than 21 years old, (b) datasets must compare at least two AMI individuals versus healthy controls or contain at least two AMI individuals, (c) samples must be derived from blood components and contain a tissue label, and (d) samples were collected within three days of AMI (Fig. 1 steps 1-2). We extracted the following information from each dataset: (1) GEO accession numbers, (2) blood source, (3) number of AMI and control individuals, (4) sex (if provided) and (5) microarray probe IDs and expression values. Information about the datasets is summarized in Table S1.

**Figure 1.**
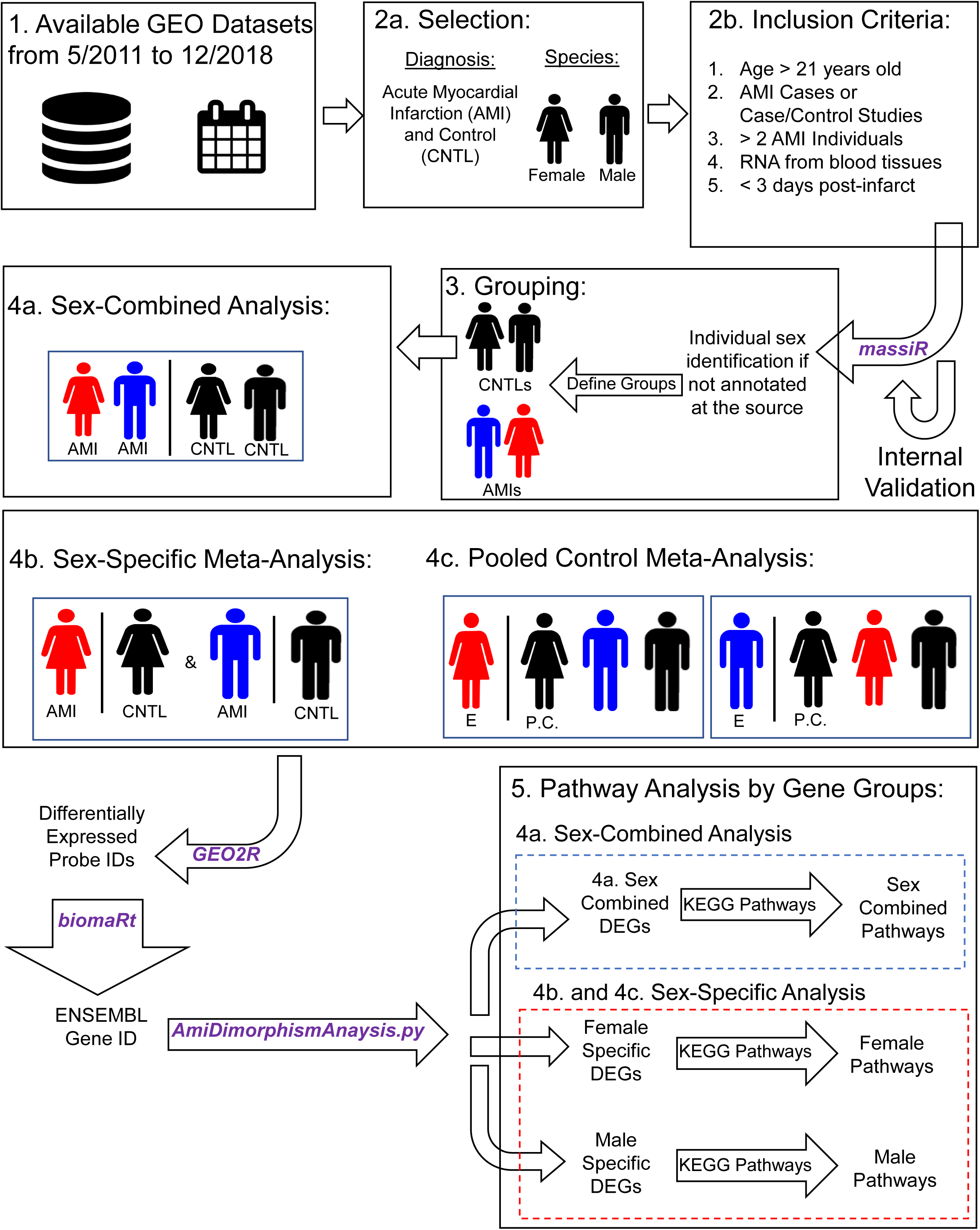
Representative schematic of study design.

### Sex identification

To perform a sex-specific analysis of gene expression, sex needed to be adjudicated to each included sample. Since most publicly available datasets did not report the sample sex, we utilized the published analysis tool, *massiR* [10], to identify the samples as a male or female. *massiR* is an R-Programming library that is designed to characterize the biological sex of samples based on counts for Y-chromosomal expression arrays. The *GEOquery* (Version 2.54.1) R-Programming library was used to input the dataset series matrix file(s) into *massiR* (Version 1.22.0). Within the *massiR* library, male samples were denoted by being above the 4^th^ quantile (threshold=4) in the *massi.select.out* command. We internally validated *massiR* by analyzing dataset GSE49925, a dataset with published sex identification, and comparing its output to the published sexes. Sex-identified individual samples within each dataset were then arranged into four groups: males with AMIs (case males), females with AMIs (case females), control males, and control females (Fig. 1-3).

**Figure 2.**
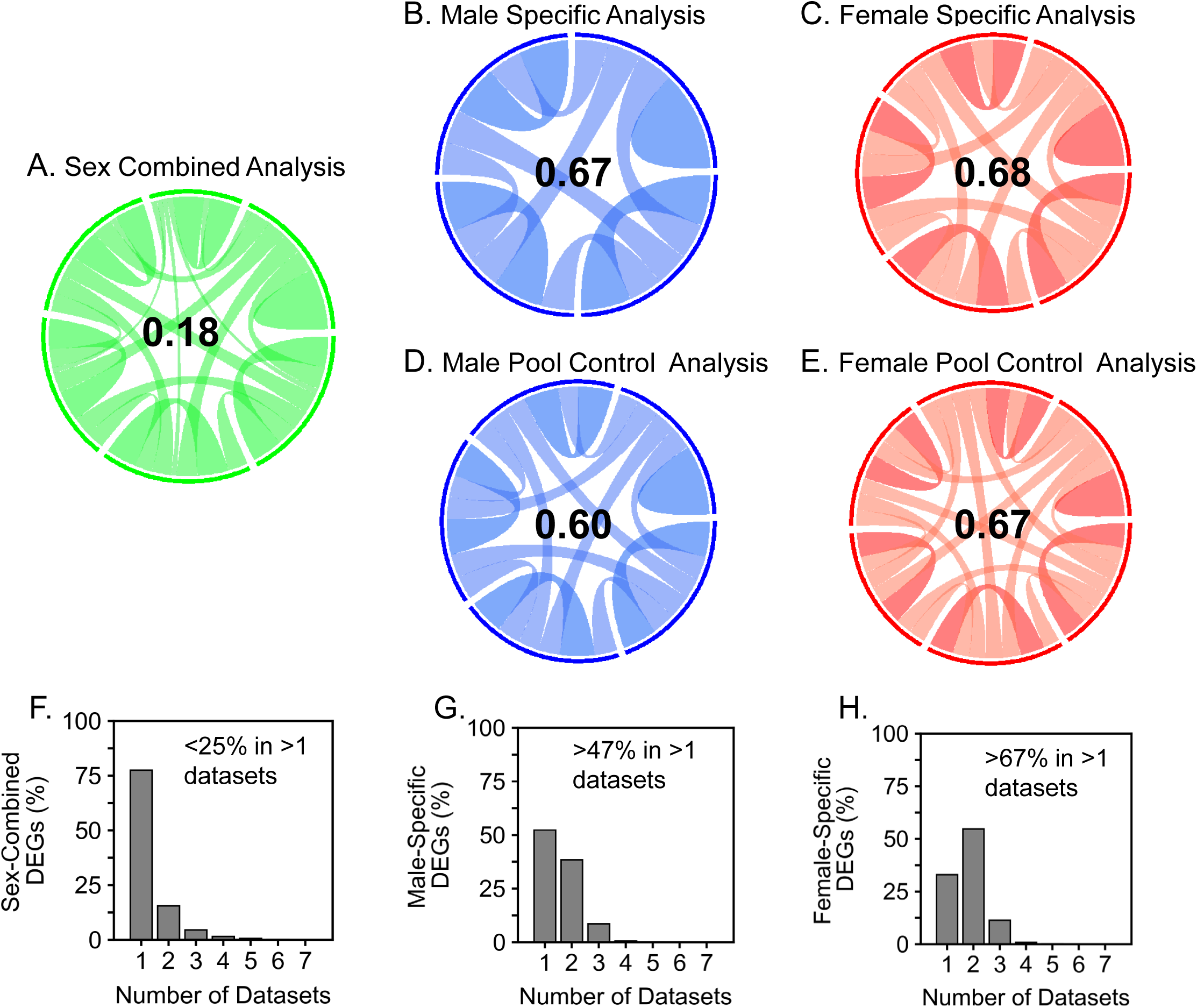
A-E) Circos plots showing Morisitia-Horn Similarity Index between datasets used in each analysis. Median index values shown. F-H) Bar graph showing the percentage of genes identified in the number of datasets based on sex-combined vs. sex-specific analysis.

**Figure 3.**
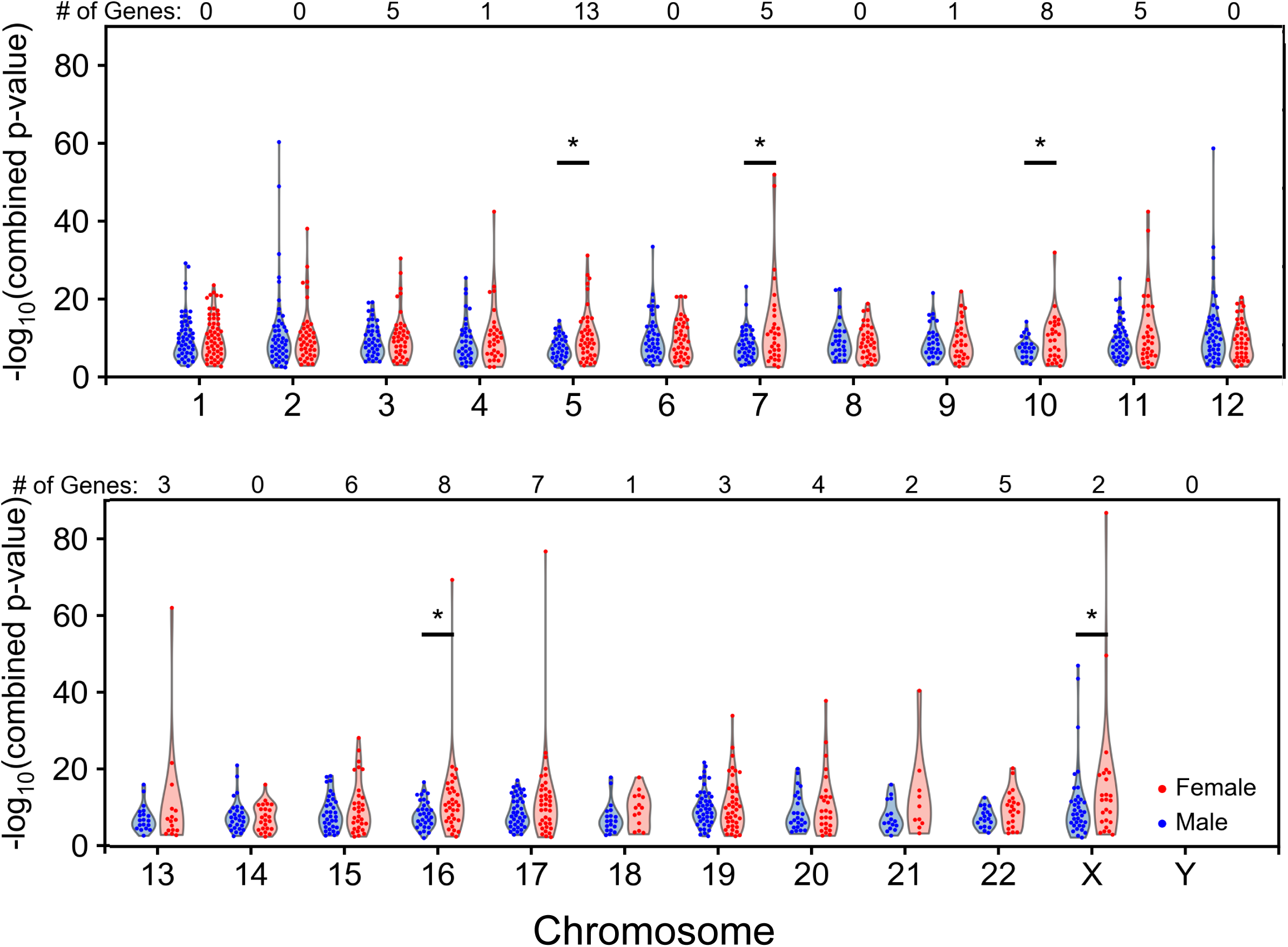
Swarmplot over violin plot depict the distribution of –log_10_ for the combined p-values for prioritize genes stemming from the sex-specific (Panel 4.b, Fig 1) and pool control analysis (Panel 4.c., Fig 1). The p-values are then mapped to their respective chromosome per sex (red= female, blue= male). *denotes the chromosomes with significantly different distributions (p<0.05, Mann-Whittney U Test) when comparing the male to the female distributions. The per-chromosome number of genes on top of the frame corresponds to the number of female-specific genes with a combined p-value < than the 99^th^ percentile of the male-specific p-values mapped to the same chromosome.

### Differential gene expression analysis

To identify differentially expressed genes (DEGs), we used the normalized gene expression data from GEO. Lists of differentially expressed microarray probe IDs were generated through the publicly available GEO2R software. We pre-specified three analysis strategies for this study: a) sex-combined, b) sex-specific and c) sex-specific pooled-control analyses. The sex-combined analysis tested all AMI cases vs. all controls (CNTLs) in a dataset (Fig. 1-4A) which yield one list of DEG per dataset. Sex-specific analysis tested AMI from a specific sex against same sex controls or against pooled controls (Fig 4B-C) which yield four additional lists of DEG per dataset.

**Figure 4.**
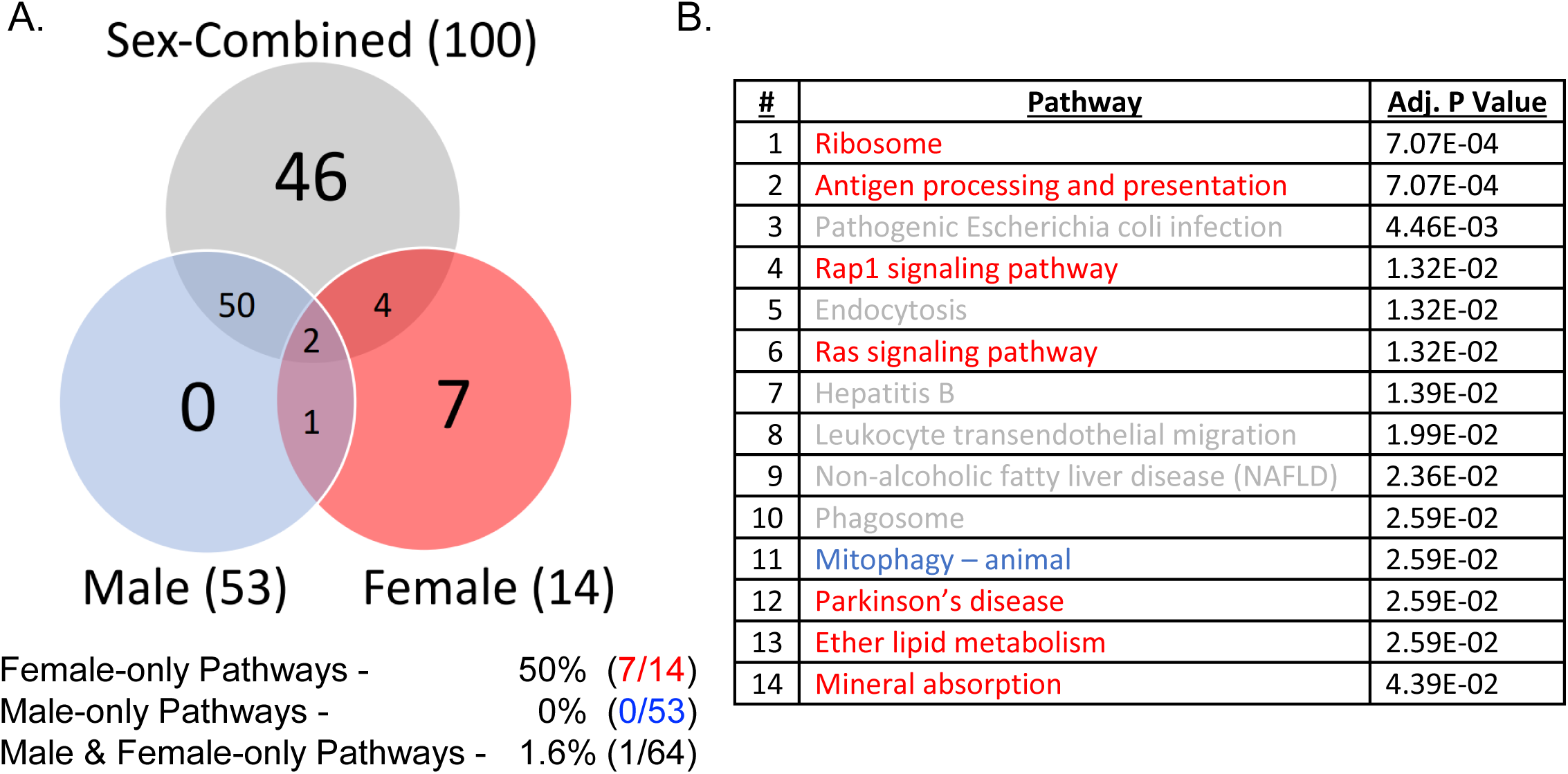
A) Venn diagram of KEGG pathways identified in sex-combined and sex-specific analyses. B) Table of pathways identified in the female-specific analysis with corresponding FDR at a threshold of <0.05 (adjusted p value). Pathways are color coordinated: grey: also appear in sex-combined analysis, blue: also appear in male-specific analysis, red: appear in the female-specific analysis only.

### Meta-Analysis of DEG

Entire lists, in both the sex-combined and sex-specific analysis, were downloaded as a tab-delimited file and written to a Microsoft Excel file by the R-programming library *writexl* (Version 1.2). Once saved in their respective excel files, the probe identities (PIDs) were converted to ENSEMBL Gene IDs by the R-programming library *biomaRt* (Version 2.42.0, Fig. 1). Not all PIDs mapped to ENSEMBL Gene IDs. This was a result of a few reasons: some PIDs mapped to pseudogenes, other PIDs were controls of the expression array experiment, etc. The mapped PIDS to ENSEMBL Gene IDs were written to excel files by *writexl*. In excel, the *VLOOKUP* function was used to pair the PIDs from GEO2R with the ENSEMBL Gene IDs from *biomaRt*. Each organized ENSEMBL Gene ID list was then copied into a new tab-delimited file for downstream analysis.

To stratify the lists of DEGs for meta-analysis, we used an in-house program using *called*, *AmiDimorphismAnalysis.py*, which utilized *sets* (Python 3.7, source code in Supplements). The program created unique sets of ENSEMBL Gene IDs from the top 1000 DEGs, ranked by p-value, that were generated from every analysis from each dataset. In the Sex-Combined, a new *set* was created by taking the union of all ENSEMBL Gene ID case v. control *sets.* In the sex-specific analysis, new *sets* were created by taking the union of ENSEMBL Gene IDs based on the comparison used to generate them. For example, a new *set* for the Case Female v. Control Female comparisons was created by taking the union of all the individual Case Female v. Control Female lists of ENSEMBL Gene IDs. Similar steps were taken for the other three groups previously mentioned. New *sets* were then created for each sex by taking the intersection of the Case v. Control and Case v. Pooled Control *sets.* For example, a new *Male Set* was created by taking the intersection of the Case Male v. Control Male *set* and Case Male v. Pooled Control *set*. Similar steps were taken to create a *Female Set*. A *Male Sex-specific Set* was created by “subtracting” the *Female Set from the Male Set.* Similar steps were taken to create a *Female Sex-specific Set*.

### Pathways Analysis

*Sets* generated by *AmiDimorphismAnalysis.py* were then run through another in-house program, *amiCleanUp.py* (Python 3.7), to create lists that formatted each term in the *sets* with its own line and remove any additional punctuation (commas, parentheses, etc.). Lists were then copied into NetworkAnalyst - Gene List Input using the following conditions: *Organism: H. Sapiens (human) and ID Type: ENSEMBL Gene ID.* Under *Assorted Visual Analytics* lists were examined by *Venn Diagram.* From the *Venn Diagram*, the space representing a singular dataset was selected. Enrichment analysis was performed through the KEGG database and top pathways (FDR<0.05) were analyzed. Similar steps were taken for every other list.

## RESULTS

### Sex-Specific Analysis Identified Novel Genes Not Identified in the Sex-Combined Analysis

To assess transcriptomic profiles of patients who recently experienced AMI, we turned to the GEO database to collect publicly available datasets from patients with acute myocardial infarction and yielded fourteen data sets (Table S1). After applying our filters (Fig. 1 steps 1-2), the included dataset consisted of 5 cohorts of blood samples: platelets [11], CD146+ Circulating Endothelial Cells [12], peripheral blood mononuclear cells [13], thrombus derived white blood cells [14], and whole peripheral blood [15–17] all collected within three days of the AMI. In total, we profiled in total 743 patient samples (39% female) from seven publicly available datasets.

Since not all available cohorts had sex denoted for each individual sample, we used *massiR*, a computational tool to assign sex based on expression data, to classify samples without sex as a reported trait. To validate *massiR*, a dataset with reported sexes (GSE49925) was first analyzed. GSE49925 consisted of 219 known males and 119 known females with no sex-unknown samples. After executing *massiR* in this dataset, we correctly classified 214/219 (97.7%) male samples and 118/119 (99.2%) female samples (Table S2). With confidence in our ability to correctly identify patient sex, we moved forward to classify the 207 sex-unknown samples in our cohort. We ultimately identified our 207 sex-unknown samples as consisting of 78 females and 129 males using *massiR*.

Lists for genes of interest were generated by cumulating the top 1,000 differentially expressed genes (ranked by *p* values) from each dataset using three distinctive analysis strategies (Fig 1. Panel 4): sex-combined analysis (4a), sex-specific meta-analysis (4b), and pooled control meta-analysis (4c). Briefly, the sex-combined analysis entailed a differential expression analysis of all patients who had an AMI with those who did not, regardless of sex. The sex-specific meta-analysis entailed a differential expression analysis of women diagnosed with AMIs and those not. This analysis was done for each dataset and then pooled across all the datasets to create our sex-combined genes of interest. These A similar approach was utilized for the men. The two analyses here were not combined nor compared in any manner. The pooled control meta-analysis entailed a differential expression analysis of women diagnosed with AMI and women who did not have an AMI, men with an AMI, and men who did not have an AMI. Said plainly, the pooled control meta-analysis compared women with AMI against the rest of our cohort, regardless of category. Again, a similar approach was taken for the men with AMI. Analysis 4b and 4c were pooled within datasets and were then pooled across datasets to generate a list of our male and female-specific genes of interest.

The sex-combined analysis (4a) yielded 4,481 genes of interest and the sex-specific analyses 2,281 genes of interest in the sex-specific analyses (4b and 4c combined). Of these 2,281 genes, 1,552 (55%) were not identified in the sex-combined analysis. In the sex-specific analyses, 1,488 (53% of 2,281) genes of interest were identified by the male analysis; 1,074 genes of interest were identified by the female analysis. In addition, 11% (259/2,281) of these genes were identified in both the male and female sex-specific analyses and were subsequently removed from each gene list to focus downstream analyses on sex-specific features.

### Identification of Genes in a Sex-Specific Analysis Yields More Redundancy than Sex-Combined

Given our sex-specific and sex-combined analysis had a similar number of input genes but different number of output genes, we hypothesized about the redundancy of genes identified in the sex-specific analysis. We modelled our differential expression gene lists for each dataset as a vector and monitored their similarity using the Morisitia Horn Similarity index (14). A Morisitia Horn Similarity Index is a continuous scale ranging from 0-1 where greater values indicate greater similarity. Figures 2A-E depict chord diagrams detailing the level of similarity between datasets in each analysis with the median similarity value in the center. In the sex-combined analysis, the median similarity value for differentially expressed gene lists across datasets was 0.18 indicating a low level of similarity across datasets and less redundancy in the final gene list. In the sex-specific analysis, median similarity index values were greater than or equal to 0.6 indicating much more similarity amongst the differentially expressed gene lists. We went on to quantify the level of redundancy across analyses and show that >75% of genes identified in the sex-combined analysis came from one 1 dataset (Fig. 2F). We additionally show that >47% of genes identified in the Male-Specific Analysis were found in at least 2 datasets (Fig. 2G). Similarly, more than 67% of genes identified in the Female-Specific Analysis were identified in at least 2 datasets (Fig. 2H).

### Sex-Specific Analysis Shows Female Preferential Usage of Specific Chromosomes

We mapped our 1,488 male-specific and 1,074 female-specific genes to their respective chromosomes to understand preferential chromosomal usage between sexes. The significance values of genes identified in the analyses were combined using Fisher’s method, log-transformed, and plotted in with respect to its chromosome. We identified five chromosomes (5, 7, 10, 16, X) which significantly different distributions of significance values (Fig. 3). We noticed in each case of significantly differently utilized chromosomes, they were more significant in the female analysis. We decided to qualify the genes identified in the female analysis whose significance values were less than the 99^th^ percentile of the male significance values within each chromosome (Table 1). In total, we identified 72 genes in the female analysis that were more significant than the respective chromosome in the male analysis.

**Table 1:** List of female-specific genes per chromosome with a p-value < than the 99^th^ percentile of the male-specific p-values in the same chromosome

| Chromosome | Gene Name |
| --- | --- |
| 3 | BCL6, RPL22P1, RPL35A, AC144530.1, RPL14 |
| 4 | SNCA |
| 5 | ATP6V0E1, BTF3, HBEGF, RPS14, HINT1, IL6ST, G3BP1, RPS23, RMND5B, ELL2, TMEM173, CD74, CD14 |
| 7 | EEF1A1P6, ACTB, MIR3654, CREB5, RAC1 |
| 9 | RPL12 |
| 10 | CH25H |
| 11 | HBB, AC104389.4, SNORD14E, ENSG00000285827, MALAT1 |
| 13 | TPT1, ITM2B, POLR1D |
| 15 | HYPK, ANXA2, B2M, OAZ2, AQP9, THBS1 |
| 16 | HBA2, C16orf54, SNORA10, SNORD68, VPS35, AHSP, ADGRG3, JPT2 |
| 17 | ACTG1, MAP2K3, AC100786.1, SLC25A39, SLC4A1, AC068152.1, SEC14L1 |
| 18 | MYL12B |
| 19 | RPS16, BCL3, R3HDM4 |
| 20 | GNAS, TGIF2-C20orf24, BCL2L1, FAM210B |
| 21 | PPIAP22, ABCC13 |
| 22 | FBXO7, DDX17, CYB5R3, SF3A1, SRRD |
| 23 | RPS4X, JPX |

### Sex-Specific Analysis Identifies Novel KEGG Pathways Not Identified in the Sex-Combined Analysis

To understand how the genes we identified would impact different cellular pathways. We ran Krypto Encyclopedia of Genes and Genomes (KEGG) Pathways Analysis to identify related pathways. Our sex-combined analysis produced 100 pathways and our sex-specific analysis produced 67 pathways (FDR<0.05, Figure 4A). 53 (79%) of the 67 pathways in the sex-specific analysis were identified in the male analysis and 14 (21%) in the female analysis. In total, we identified 8 pathways in our sex-specific analysis that were not denoted in our sex-combined analysis. Interestingly, we did not identify any pathways that were unique to the male analysis but did identify 7 (88% of pathways in the sex-specific analysis) that were unique to the female analysis (FDR<0.05, Figure 4B). These seven female-specific pathways included ‘Ribosome’, ‘Antigen processing and presentation’, ‘Rap1 signaling pathway’, ‘Ras signaling pathway’, ‘Parkinson’s disease’, ‘Ether lipid metabolism’, and ‘Mineral absorption’. We additionally identified 1 pathway, ‘Mitophagy—animal’, that was shared between the female and male pathway analyses.

## DISCUSSION

Our findings underscore the necessity for studying female specific cardiovascular biology by demonstrating that when women are studied in isolation from men, previously uncharacterized genes and pathways become potential avenues for further investigation. This unravels sex-specific and shared biological pathways while underscoring a need for prospective sex-specific transcriptome and clinical outcome studies to support future treatments post-AMI. AMIs are complex and elusive events that impact male and female mortality rates differently. Regardless of nearly identical genomic sequences, transcriptomic differences between the sexes can change the regulation of certain pathways and lead to the disparity in patient phenotypes. Our goal was to perform parallel sex-specific and sex-combined analyses to determine the efficacy of sex as an experimental variable of AMI mortality. Previous studies like this have been uncommon due to the lack of publication of patient sex in GEO. Our sex-specific analysis was able to identify 2,281 DEGs, 1,552 (55%) of which, were not identified in the sex-combined analysis.

Datasets included in the study were similar to the datasets excluded in multiple respects. The RNA sources from the included datasets were mostly whole blood samples, similar to excluded datasets (Table S1) [18–23]. Most types of AMIs reported were designated as AMIs with no further specification. STEMI was noted as the most common specification of AMIs in our included and excluded datasets. Datasets could not create all four analysis comparisons because they only contained one AMI patient of a particular sex.

Our analysis yielded two interesting findings. First, the DEGs in the sex-specific analysis showed more reproducibility across datasets, adding validity to their findings and importance. The sex-combined analysis produced 4,481 unique DEGs. Over 75% of these genes occurred in only one dataset. The lack of reproducibility across datasets does not inspire confidence the genes identified are likely to be informative to the disparity in mortality rates. Reproducibility is such a strong factor of determining the validity of an experiment that journals such as *Nature* have released articles on the “reproducibility crisis” in science [24 25]. Second, 50% of the pathways identified in the female-specific analysis were not identified in the sex-combined analysis implying the dramatic loss of female signal in canonical analyses. These newly identified pathways and genes are promising outlets for further investigation and inquiry.

In summary, our discovery pipeline identified a series of novel genes and pathways to be explored and validated as factors in the etiology of the sexual dimorphism of AMI mortality rates. Our results underscore the importance of including sex as a variable of interest in future clinical studies to better understand the disparity. More research is required to validate the results shown here and develop sex-specific treatments for patients suffering from an AMI to best increase their survival. By identifying sex specific DEGs, we were able to identify several pathways that could also be informative to the disparity. Our study serves to build a bridge between genomics and clinical treatments to increase patient survival and recovery.

## Data Availability

All data will be made available upon request to the corresponding author.

## ACKNOWLEDGMENTS

We would like to thank and acknowledge the original authors of the studies involved in this meta-analysis for their diligence in their study design. We would like to also thank the original author or *massiR*, Dr. Buckman, for their additional guidance on the proper use of their tool.

## SUPPLEMENTAL MATERIALS

**Table S1:** GEO Datasets Identified in initial query and used in final analysis.

|  |  | Dataset | Source | Samples in Dataset |  |  |  | Analysis Involved In |  |  |  |
| --- | --- | --- | --- | --- | --- | --- | --- | --- | --- | --- | --- |
|  |  |  |  | # of Case Males | # of Case Females | # of Control Male | # of Control Female | Case Male v. Control Male | Case Male v. Pooled Control | Case Female v. Control Female | Case Female v. Pooled Control |
| Used in Analysis | Sex Identification Given | GSE24519 | Platelets | 20 | 10 | 84 | 84 |  | x |  | x |
|  |  | GSE49925 | Peripheral Blood | 219 | 119 |  |  |  | x |  | x |
|  | Sex Identification by *massIR* | GSE48060 | Peripheral Blood | 17 | 14 | 14 | 7 | x | x | x | x |
|  |  | GSE97320 | Peripheral Blood | 1 | 2 | 1 | 2 |  | x | x | x |
|  |  | GSE66360 | CD146+ Circulating Endothelial Cells | 38 | 11 | 20 | 30 | x | x | x | x |
|  |  | GSE62646 | PBMCs | 21 | 7 | 11 | 3 | x | x | x | x |
|  |  | GSE19339 | Thrombus derived WBCs | 3 | 1 | 3 | 1 | x | x |  | x |
| Not Used in Analysis | Incapatiable with *massIR* | GSE53211 | CD34+ Progenitor Cells | 4 Total Case Samples |  | 0 Total Control Samples |  |  |  |  |  |
|  |  | GSE61144 | Peripheral Blood | 7 Total Case Samples |  | 10 Total Control Samples |  |  |  |  |  |
|  |  | GSE31568 | Peripheral Blood | 20 Total Case Samples |  | 20 Total Control Samples |  |  |  |  |  |
|  | Incapatible with *biomaRt* | GSE123487 | Peripheral Blood | 3 | 1 | 0 | 0 |  |  |  |  |
|  |  | GSE24548 | Platelets | 4 | 0 | 64 | 64 |  |  |  |  |
|  |  | GSE29532 | Peripheral Blood | 9 | 0 | 6 | 0 |  |  |  |  |
|  | Produced too Few Significant DEGs | GSE28454 | Monocytes | 19 | 7 | Not Published At Time of Analysis |  |  |  |  |  |
|  | Totals of Datasets Used in the Anlaysis: |  |  | 319 | 164 | 133 | 127 | 4 | 7 | 4 | 7 |
|  | Totals of Datasets Not Used in the Analysis: |  |  | 74 Total Cases |  | 164 Total Controls |  |  |  |  |  |

**Table S2:** massiR validation using GSE49925.

| **GSE49925** | **<u>Truth</u>** | **<u>*massiR*</u>** | **<u>Correctly Called</u>** |
| --- | --- | --- | --- |
| **Males** | 219 | 214 | 97.7% |
| **Females** | 119 | 118 | 99.2% |

## AUTHOR CONTRIBUTIONS

Formal Analysis: A.S.

Methodology and Investigation: A.S., P.P., E.V., E.C.T., T.B.M., J.E.L.

Resources: K.A., J.E.L.

Writing – Original Draft: A.S., P.P., J.E.L.

Funding Acquisition: J.E.L.

Supervision: T.B.M., J.E.L.

## REFERENCES

1. Ober C, Loisel DA, Gilad Y. Sex-specific genetic architecture of human disease. Nat Rev Genet 2008;9(12):911–22 doi: 10.1038/nrg2415.

2. Gautam Y, Afanador Y, Abebe T, Lopez JE, Mersha TB. Genome-wide analysis revealed sex-specific gene expression in asthmatics. Hum Mol Genet 2019;28(15):2600–14 doi: 10.1093/hmg/ddz074.

3. Manzo-Silberman S, Hawranek M, Banerjee S, et al. Call to action for acute myocardial infarction in women: international multi-disciplinary practical roadmap. Eur Heart J Open 2024;4(6):oeae087 doi: 10.1093/ehjopen/oeae087 [published Online First: 20241012].

4. Mosca L, Barrett-Connor E, Wenger NK. Sex/gender differences in cardiovascular disease prevention: what a difference a decade makes. Circulation 2011;124(19):2145–54 doi: 10.1161/CIRCULATIONAHA.110.968792.

5. Martin SS, Aday AW, Almarzooq ZI, et al. 2024 Heart Disease and Stroke Statistics: A Report of US and Global Data From the American Heart Association. Circulation 2024;149(8):e347–e913 doi: 10.1161/CIR.0000000000001209 [published Online First: 20240124].

6. Mehta LS, Beckie TM, DeVon HA, et al. Acute Myocardial Infarction in Women: A Scientific Statement From the American Heart Association. Circulation 2016;133(9):916–47 doi: 10.1161/CIR.0000000000000351 [published Online First: 20160125].

7. Dungan JR, Qin X, Gregory SG, et al. Sex-dimorphic gene effects on survival outcomes in people with coronary artery disease. Am Heart J Plus 2022;17 doi: 10.1016/j.ahjo.2022.100152 [published Online First: 20220614].

8. Huaqiang Zhou KY, Shaowei Gao, Yuanzhe Zhang, Xiaoyue Wei, Zeting Qiu, Si Li, Qinchang Chen, Yiyan Song, Wulin Tan, Zhongxing Wang. Identification of differentially expressed genes between male and female patients with acute myocardial infarction based on microarray data. International Journal of Clinical and Experimental Medicine 2019 doi: 2019;12(3):2456–2467.

9. Barrett T, Wilhite SE, Ledoux P, et al. NCBI GEO: archive for functional genomics data sets— update. Nucleic acids research 2012;41(D1):D991–D95.

10. Buckberry S, Bent SJ, Bianco-Miotto T, Roberts CT. massiR: a method for predicting the sex of samples in gene expression microarray datasets. Bioinformatics 2014;30(14):2084–5 doi: 10.1093/bioinformatics/btu161 [published Online First: 20140322].

11. Bellin M NL, De Pittà C, Viganò A, Vallanti G, Vasso M, Vozzi D, Cristell N, Cianflone D, Gelfi C, Maseri A, Lanfranchi G. Gene expression profiling of patients affected by first acute myocardial infarction (FAMI). GEO, 2017.

12. Muse ED, Kramer ER, Wang H, et al. A Whole Blood Molecular Signature for Acute Myocardial Infarction. Sci Rep 2017;7(1):12268 doi: 10.1038/s41598-017-12166-0 [published Online First: 20170925].

13. Kiliszek M, Burzynska B, Michalak M, et al. Altered gene expression pattern in peripheral blood mononuclear cells in patients with acute myocardial infarction. PLoS One 2012;7(11):e50054 doi: 10.1371/journal.pone.0050054 [published Online First: 20121121].

14. Fu H, Vadalia N, Xue ER, et al. Thrombus leukocytes exhibit more endothelial cell-specific angiogenic markers than peripheral blood leukocytes do in acute coronary syndrome patients, suggesting a possibility of trans-differentiation: a comprehensive database mining study. J Hematol Oncol 2017;10(1):74 doi: 10.1186/s13045-017-0440-0 [published Online First: 20170323].

15. Kim J, Ghasemzadeh N, Eapen DJ, et al. Gene expression profiles associated with acute myocardial infarction and risk of cardiovascular death. Genome Med 2014;6(5):40 doi: 10.1186/gm560 [published Online First: 20140530].

16. F M. Differential gene expression profiles in peripheral blood in Northeast Chinese Han people with acute myocardial infarction: GEO, 2017.

17. Suresh R, Li X, Chiriac A, et al. Transcriptome from circulating cells suggests dysregulated pathways associated with long-term recurrent events following first-time myocardial infarction. J Mol Cell Cardiol 2014;74:13–21 doi: 10.1016/j.yjmcc.2014.04.017 [published Online First: 20140504].

18. Park HJ, Noh JH, Eun JW, et al. Assessment and diagnostic relevance of novel serum biomarkers for early decision of ST-elevation myocardial infarction. Oncotarget 2015;6(15):12970–83 doi: 10.18632/oncotarget.4001.

19. Keller A, Leidinger P, Bauer A, et al. Toward the blood-borne miRNome of human diseases. Nat Methods 2011;8(10):841–3 doi: 10.1038/nmeth.1682 [published Online First: 20110904].

20. Li Y, He XN, Li C, Gong L, Liu M. Identification of Candidate Genes and MicroRNAs for Acute Myocardial Infarction by Weighted Gene Coexpression Network Analysis. Biomed Res Int 2019;2019:5742608 doi: 10.1155/2019/5742608 [published Online First: 20190211].

21. Bellin M NL, De Pittà C, Viganò A, Vallanti G, Vasso M, Vozzi D, Cristell N, Cianflone D, Gelfi C, Maseri A, Lanfranchi G. microRNA expression profiling of patients affected by first acute myocardial infarction (FAMI): GEO, 2017.

22. Silbiger VN, Luchessi AD, Hirata RD, et al. Novel genes detected by transcriptional profiling from whole-blood cells in patients with early onset of acute coronary syndrome. Clin Chim Acta 2013;421:184–90 doi: 10.1016/j.cca.2013.03.011 [published Online First: 20130325].

23. Maouche S SH, Aherrahrou Z, Belz S, Brocheton J, Proust C, Tennstedt S, Ammerlahn H, Riedel C, Stritzke J, Kurowski V, Rice KM, Goodall AH, Hengstenberg C, Ouwehand WH, Samani NJ, Cambien F, Erdmann J, Schunkert H, Diemert P. Temporal transcriptional changes in human monocytes following acute myocardial infarction: The GerMIFs monocyte expression study: GEO, 2012.

24. Adam D. What reproducibility crisis? New research protocol yields ultra-high replication rate. Nature 2023;623(7987):467-68 doi: 10.1038/d41586-023-03486-5.

25. Baker M. 1,500 scientists lift the lid on reproducibility. Nature 2016;533(7604):452–4 doi: 10.1038/533452a.

